# The computerised PROTECT Cognitive Test System is sensitive to dementia and correlates with Alzheimer-related blood biomarkers

**DOI:** 10.1101/2025.10.13.25337742

**Authors:** Anne Corbett, Abbie Palmer, Millie Sander, Christine Davis, Kate Stych, Mary O’Leary, Jon Huntley, Dag Aarsland, Nicolas Castellanos-Perilla, Felipe Botero-Rodriguez, Gerard Griffioen, Mieke Nuytten, Nicholas Ashton, Hanna Huber, Adam Hampshire, Clive Ballard, the REACTIVE Investigator Group

**Affiliations:** University of Exeter Medical School, University of Exeter, Exeter, UK; Centre for Age-Related Medicine (SESAM), University Hospital, Stavanger, Norway; Department of Neurobiology, Care Sciences and Society, Karolinska Institutet, Stockholm, Sweden; Department of Clinical Medicine, University of Bergen, Bergen, Norway; SynaptIA, Bogotá, Colombia; remynd, Heverlee, Belgium; University of Gothenberg, Gothenberg, Sweden; German Center of Neurodegenerative Diseases, Bonn, Germany; Department of Neuroimaging, King’s College London, London, UK

## Abstract

Early detection of cognitive impairment is essential for timely diagnosis and to support recruitment of pre-clinical patients into trials of disease-modifying therapies. Current clinically used tools have limited sensitivity for early cognitive deficit and require in-person assessments. Computerised assessment systems offer a means of addressing these challenges with scalable self-administered domain-specific assessments for use in the community and primary care. This study presents further validation of the PROTECT Cognitive Test System of eight assessments of memory, attention and executive function in 113 patients with dementia and 9197 cognitively healthy controls. PROTECT shows robust separation of dementia and non-dementia patients in all cognitive tests and domains (P<0.001), and a Receiver Operator Characteristic analysis also shows excellent discriminative ability (>0.9), >90% sensitivity and >89% specificity in all domains. The system also correlates strongly with the plasma biomarker p-tau217 (P<0.001, AUC 0.885) and with the MoCA (P=0.006). The self-administered online delivery means it offers a means of triaging large numbers of patients for early diagnostics and trials and the strength of correlation with an AD biomarker is very favourable compared to other assessment instruments.

## Introduction

There are one million people with dementia in the UK, with an annual cost of £42billion^1^. Mild Cognitive Impairment (MCI) affects up to 15% of adults over 50 with 10% developing dementia annually^2^. Despite the importance of early diagnosis and support, 99% of people with MCI are not in contact with specialist health services and most wait until they reach a crisis before seeking help^3^. Emerging disease-modifying therapies have the potential to change the landscape of dementia treatment and early prevention but they will require an efficient patient funnel in the community and primary care to support early diagnosis in specialist settings^4–6^. Clinical trials also rely on efficient identification of people with early cognitive decline since these are the primary target patient groups for emerging treatments^7^. Existing health services models do not have capacity or suitable assessment tools to support large-scale patient triage for diagnosis or trials. Furthermore, the absence of an efficient patient funnel hampers recruitment to large preventative and early-intervention trials.

Diagnosis of dementia currently involves a combination of clinical history taking, neuropsychological testing, biomarker testing and neuroimaging^8^. However, this process is intensive, invasive and expensive and requires efficient triage to create an effective diagnostic pathway. Traditional paper and pencil assessments of cognition, such as the Mini Mental State Examination, Alzheimer’s Disease Assessment Scale-Cognitive Subscale (ADAS-Cog) and the Montreal Cognitive Assessment (MoCA) are easy to use but have learning effects, have limited ability to detect changes in specific cognitive domains, and rely on a trained health professional^9^. As a result, these tools are not suitable for scalable cognitive assessment. Computerised cognitive testing and remote assessment offers a means to improve on the current status quo for brain health assessment in both clinical and research settings. Computerised tests offer consistent, unbiased, highly sensitive recording of cognitive function, which is key to diagnosis and early detection of cognitive decline^10–12^ as well as for quantifying treatment outcomes in clinical trials^13^. Importantly, computerised tests raise the potential for remote, unsupervised assessment which makes them scalable. Over 30 digital cognitive test systems exist, and the field is rapidly growing, but supporting evidence is hampered by a lack of longitudinal data validation and clinical validation^14^.

The PROTECT Cognitive Test System is a suite of computerised neuropsychological assessments that has been developed and validated as part of a portfolio of research over the last ten years. It has undergone full computer systems and data validation in the PROTECT-UK cohort of 50,000 participants, with over 2.5million assessments completed^15^. The system has shown ability to detect statistically meaningful age-related cognitive decline, demonstrating concurrent validity with established FDA-approved criteria for early cognitive impairment, and sensitivity to trajectory of cognitive decline in healthy and impaired individuals^16^. It has been used to measure cognitive change in several completed and ongoing clinical trials and ageing cohorts, which provides the opportunity to further validate its sensitivity and accuracy in detecting clinically-meaningful cognitive status^17–21^.

There is also an exciting opportunity to combine computerised neuropsychology with rapidly emerging blood biomarkers for neurodegeneration. Blood biomarkers are becoming well established for AD risk detection and are now being adopted into clinical practice^22^. The most robust blood biomarker is phosphorylated tau at amino acid 217 (p-tau217), which shows highest accuracy for detection of amyloid pathology ^23,24 25,26^. The combination of blood biomarkers with computerised neuropsychology offers the potential for scalable monitoring and assessment of early cognitive decline and dementia risk which could be embedded in primary care and support patient funnels through to specialist services or clinical trials.

This study presents a combined analysis of data from healthy and clinical dementia cohorts to further validate the PROTECT Cognitive Test System for detection of dementia and concurrent validity clinical dementia assessment and blood biomarkers.

## Methods

### Study Design

This is a cross-sectional study to establish the concurrent validity, sensitivity and specificity of the PROTECT Cognitive Test System in detecting dementia. The study sought to determine its ability to differentiate between individuals with and without dementia and to determine concurrent validity with neurodegenerative blood biomarkers by analysing data derived from patients in five international sites. Ethical approval was obtained from the (NorthWest Greater Manchester East NHS Research Ethics Committee Ref: 24/NW/0151).

### Participants

All participants were aged between 54 and 90 years old, had capacity to consent and were able to use a computer or touchscreen device. Participants were excluded if they had a visual impairment, neurological condition, stroke affecting upper limbs or any physical disability impairing the use of a device. Dementia patients fulfilled criteria for mild to moderate dementia based on the National Institute of Neurological and Communicative Disorders and Stroke and Alzheimer’s Disease and Related Disorders Association (NINCDS-ADRDA) criteria with impairment in at least two cognitive domains with impact on functionality. PROTECT-UK participants were healthy volunteers with no diagnosis of dementia, residing in the UK.

### Recruitment and consent

Participants with dementia were recruited from primary and secondary clinical sites in the UK, Spain, Norway and the Netherlands^13,16,17,20,21^. Patients were pre-screened for eligibility criteria and provided written informed consent. Cognitively healthy participants in the PROTECT-study were recruited across the UK as part of the study open recruitment phase between 2014 and 2024. Registration and informed consent was given online through an ethically approved digital process, including consent for use of data for research purposes. All participants provided demographic information at registration.

### Cognitive Assessments

All participants completed the PROTECT Cognitive Test System on a computer or touchscreen device. The system consists of eight individual tests for working memory, episodic memory, executive function and attention. The tests are Paired Associate Learning, Digit Span, Self-Ordered Search, Simple Reaction Time, Choice Reaction Time, Digit Vigilance, Delayed Picture Recognition and Verbal Reasoning. Description of these tests and their outputs are shown in Supplementary Table 1. Participants completed a brief practice session of each test in sequence prior to completing a full test session in order to reduce practice effects due to orientation to the system. Tests run in a pre-scheduled sequence and start automatically to ensure consistency of delivery. Participants in the dementia cohort completed the test system under supervision from the medical team. Staff were available to support in the event of a technical or medical issue but did not assist in completion of the tests themselves. Dementia cohort participants also completed the Montreal Cognitive Assessment (MoCA), as per published standard protocols, with a trained medical staff team member.

**Table 1.** Participant characteristics.

|  | N | Age |  | Sex |
| --- | --- | --- | --- | --- |
|  |  | Range | Mean (SD) | Female n (%) |
| **Dementia patients** | 113 | 55 - 89 | 73.77 (8.33) | 45 (39) |
| **Healthy comparators** | 9197 | 55-89 | 64.29 (6.00) | 6529 (71) |

Participants in the PROTECT-UK cohort completed the Cognitive Test System annually as part of the longitudinal data collection protocol. Participants accessed the system from the PROTECT-UK study online dashboard and completed the tests unsupervised at home.

### Blood Biomarker Collection

A subset of participants provided a venous blood sample by venipuncture for biomarker analysis at a clinic visit, taken by a trained staff member. Prior to analysis, plasma samples were thawed, vortexed for 30 seconds at 2,000 rpm and centrifuged at 4,000 g and 20°C for 10 minutes. Biomarker analysis was completed by Single molecule array (Simoa) technology using a 2X dilution protocol according to manufacturer’s instructions (ALZpath Simoa^®^ p-tau217 v2 Assay,) (Quanterix, Billerica, MA, USA) to detect calibrated levels of p-tau217.

### Analysis

Descriptive statistics were generated for each cohort. Cognitive data from the Cognitive Test System were extracted from the electronic database for each cohort and combined for analysis, with each test variable represented and categorised by cognitive domain. Composite cognitive scores were calculated according to published protocols. An age-matched comparator dataset was created from the PROTECT-UK cohort. Concurrent validity with the MoCA and blood biomarker levels were analysed using the Pearson’s R and Spearman’s R tests respectively. Comparative analysis with the PROTECT-UK dataset was completed by t-test. Data were categorised as dementia or healthy based on the source cohort, Area Under the Curve (AUC) of receiver operating characteristics (ROC) was computed, and the sensitivity and specificity was generated. Analyses were completed in SPSS Software (Version 31) and R Studio (Version 4.4.3).

## Results

### Cohort Characteristics

This study included data from 113 patients with dementia and 9197 healthy participants in the age-matched dataset, with age range from 54 – 90 (Table 1). Of the 113 dementia patients, three did not complete the full cognitive test system. Full data was available for all other patients.

### Differentiation between dementia and healthy patients

Comparative analysis of cognitive performance between the dementia and healthy cohorts showed significant difference between all cognitive test outputs for memory, attention and executive function including all composite measures (Table 2).

**Table 2.**
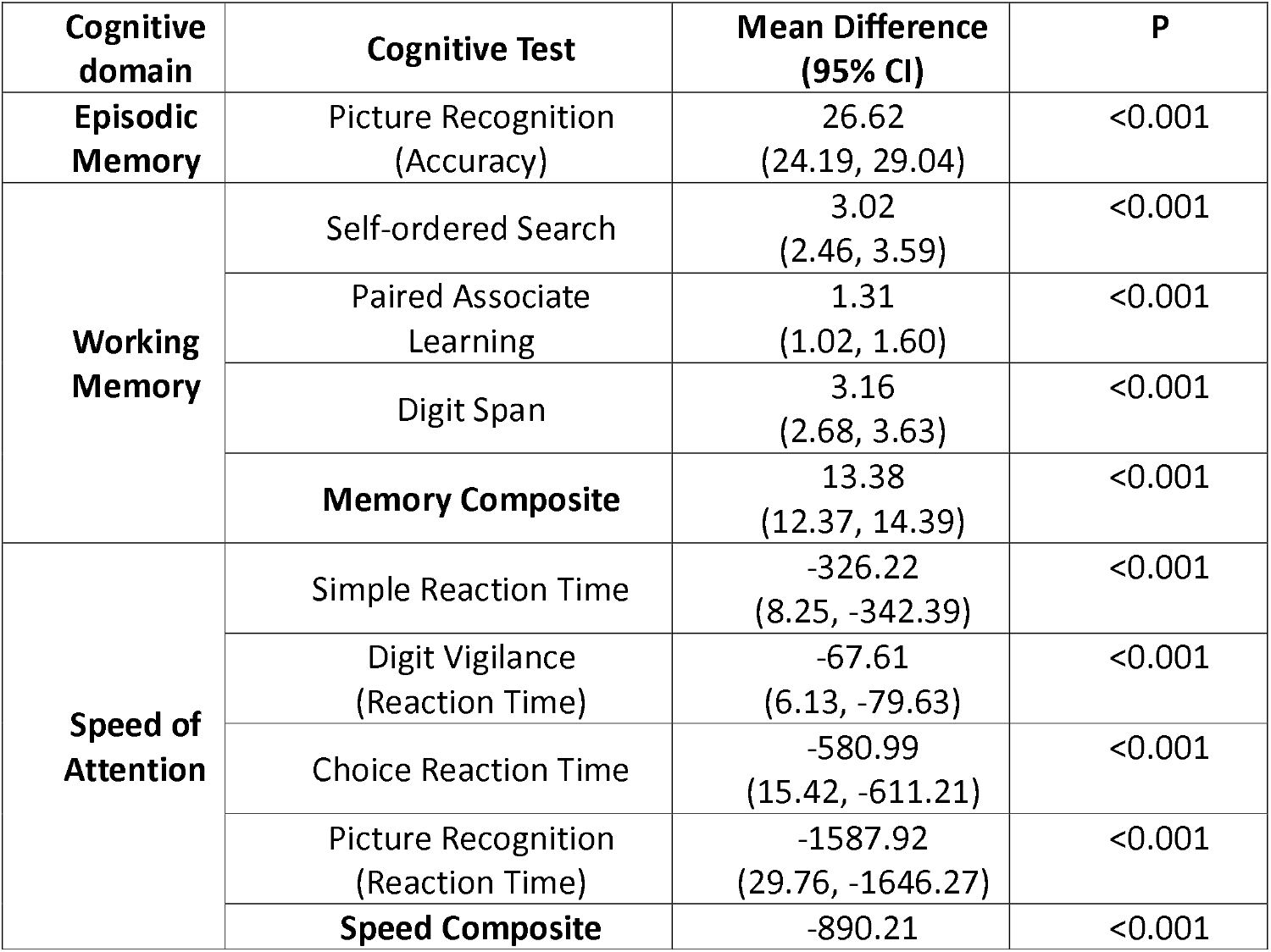
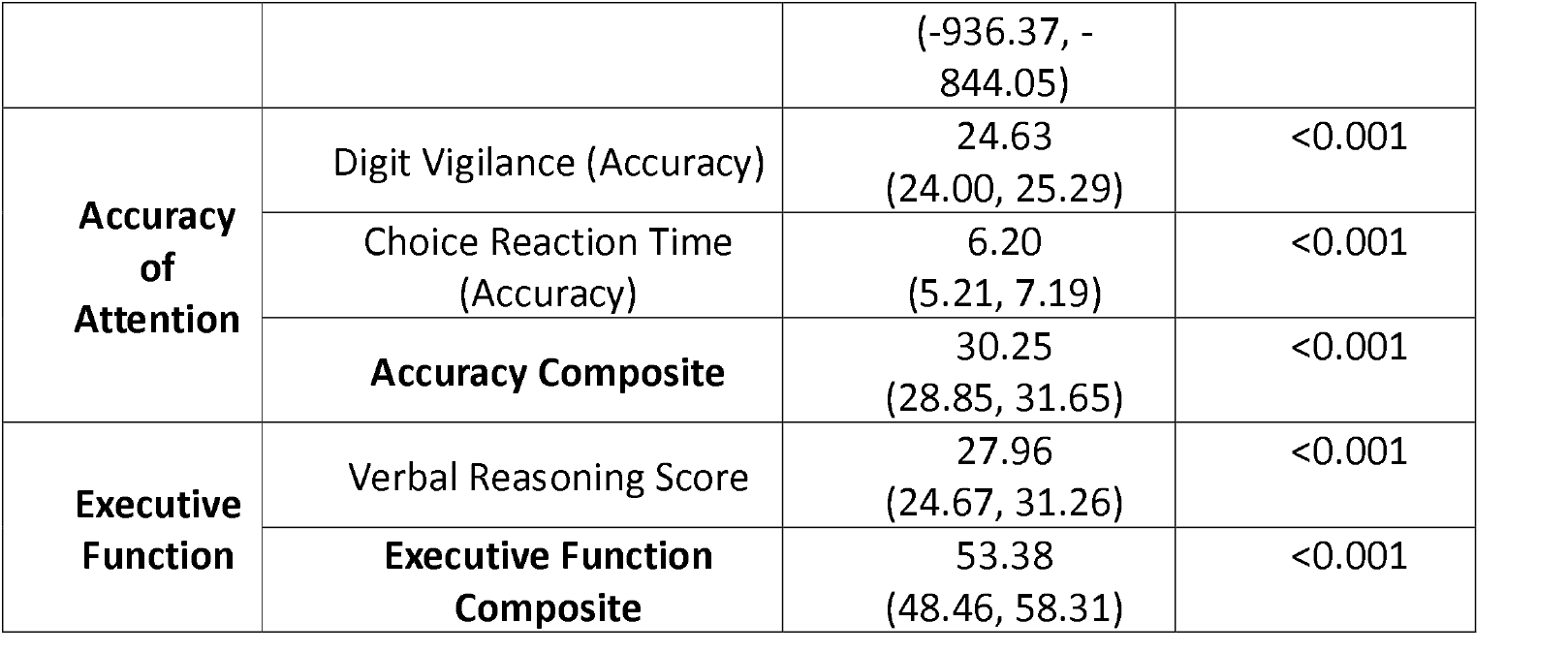
Comparison of cognitive performance between dementia cohorts and healthy counterparts shows significant difference across all cognitive tests and domains (n=9250)

### ROC analysis

ROC analysis comparing dementia and healthy cohort data from the PROTECT Cognitive Test System showed significant Area Under the Curve values for each cognitive domain, reaching 90% sensitivity and over 89% specificity for all domains (P<0.001) (Table 3; Figure 1).

**Table 3.** Receiver Operator Characteristic output shows robust discrimination between dementia and healthy cohorts for all cognitive domains (n=9250)

| Cognitive Domain | AUC<br>(95% CI) | P | Sensitivity | Specificity | Optimal<br>cut-off<br>score |
| --- | --- | --- | --- | --- | --- |
| **Memory** | 0.922<br>(0.919, 0.984) | <0.001 | 0.905 | 0.900 | 73.75% |
| **Speed of Attention**<br>**(Reaction Time)** | 0.966<br>(0.934, 0.990) | <0.001 | 0.911 | 0.889 | 1422.75ms |
| **Accuracy of**<br>**Attention**<br>**(Accuracy)** | 0.960<br>(0.924, 0.997) | <0.001 | 0.918 | 0.900 | 94.45% |
| **Executive Function** | 0.957<br>(0.921, 0.995) | <0.001 | 0.902 | 0.903 | 23.500 |

**Figure 1.**
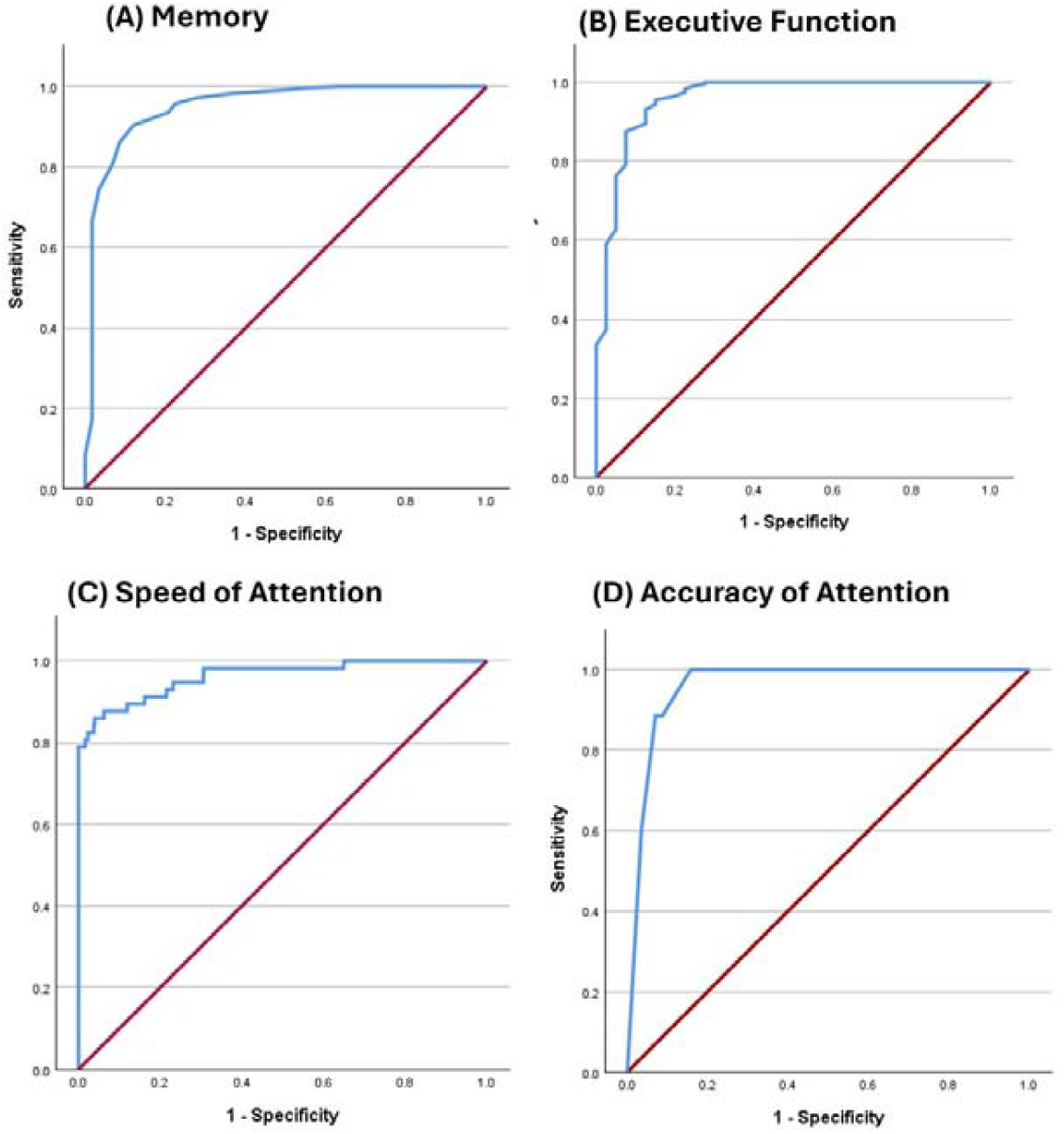
ROC curves for memory, executive function, speed of attention and accuracy of attention. ROC analysis shows robust discrimination between dementia patients and healthy participants on the cognitive domains of Memory (A), Executive Function (B), Speed of Attention (C) and Accuracy of Attention (D) tested by the PROTECT Cognitive Test System

### Concurrent validity with the MoCA

MoCA score data was available for 59 participants. Correlation analysis showed significant correlation of the PROTECT Cognitive Test System outputs with MoCA score in executive function (P=0.043) and accuracy of attention (P = 0.006), including the composite scores, and significant correlation in tests of episodic memory (P=0.042) and working memory except for Paired Associate Learning (Memory composite P=0.020). In Speed of Attention there were significant correlations in reaction time in the Digit Vigilance (P=0.021) and Picture Recognition (P=0.010) tests but not in the Simple or Choice reaction time tests or in the Speed of Attention composite. Full analysis outputs are shown in Table 4.

**Table 4.**
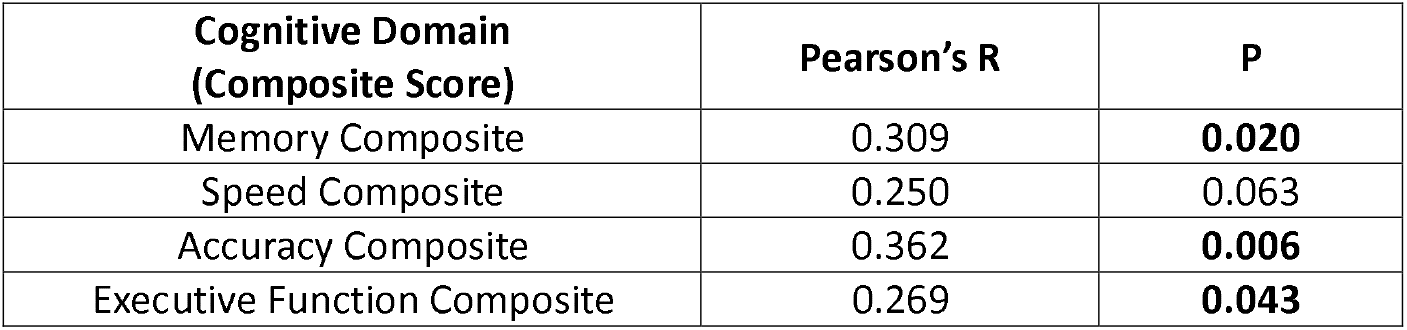
Concurrent validity of PROTECT Cognitive Test variables with MoCA score (n=59)

| Cognitive Domain<br>(Composite Score) | Pearson's R | P |
| --- | --- | --- |
| Memory Composite | 0.309 | **0.020** |
| Speed Composite | 0.250 | 0.063 |
| Accuracy Composite | 0.362 | **0.006** |
| Executive Function Composite | 0.269 | **0.043** |

### Correlation with venous blood biomarkers for neurodegeneration

Blood biomarker data was available from 77 participants across the PROTECT (n = 31) and Dementia (n = 46) cohorts. Analysis showed significant correlations between p-tau217 and all tests of episodic and working memory (r = 0.44-0.55). P-tau217 also showed significant correlation with executive function, speed of attention and the Digit Vigilance test for accuracy of attention but not the Choice Reaction Time accuracy measure or the accuracy of attention composite (Table 5).

**Table 5.** Correlation of the PROTECT Cognitive Test System with blood biomarkers p-tau217 (n=77)

| Cognitive domain | Cognitive Test | Plasma p-tau217 |  |
| --- | --- | --- | --- |
|  |  | Spearman's R | P |
| Episodic Memory | Picture Recognition (Accuracy) | -0.547 | <0.001 |
| Working Memory | Self-ordered Search | -0.466 | <0.001 |
|  | Paired Associate Learning | -0.443 | <0.001 |
|  | Digit Span | -0.491 | <0.001 |
|  | Memory Composite | -0.667 | <0.001 |
| Speed of Attention | Simple Reaction Time | 0.388 | 0.001 |
|  | Digit Vigilance (Reaction Time) | 0.374 | 0.003 |
|  | Choice Reaction Time | 0.414 | <0.001 |
|  | Picture Recognition (Reaction Time) | 0.354 | 0.004 |
|  | Speed Composite | 0.455 | <0.001 |
| Accuracy of Attention | Digit Vigilance (Accuracy) | 0.641 | 0.002 |
|  | Choice Reaction Time (Accuracy) | 0.102 | 0.424 |
|  | Accuracy Composite | -0.252 | 0.269 |
| Executive Function | Verbal Reasoning Score | -0.521 | <0.001 |
|  | Executive Function Composite | -0.506 | <0.001 |

ROC analysis showed robust discrimination of p-tau217 biomarker levels (AUC = 0.885, P<0.001, 95%CI 0.775-0.979; Figure 2) between dementia and non-dementia groups, reaching 92% sensitivity and 79% specificity.

**Figure 2.**
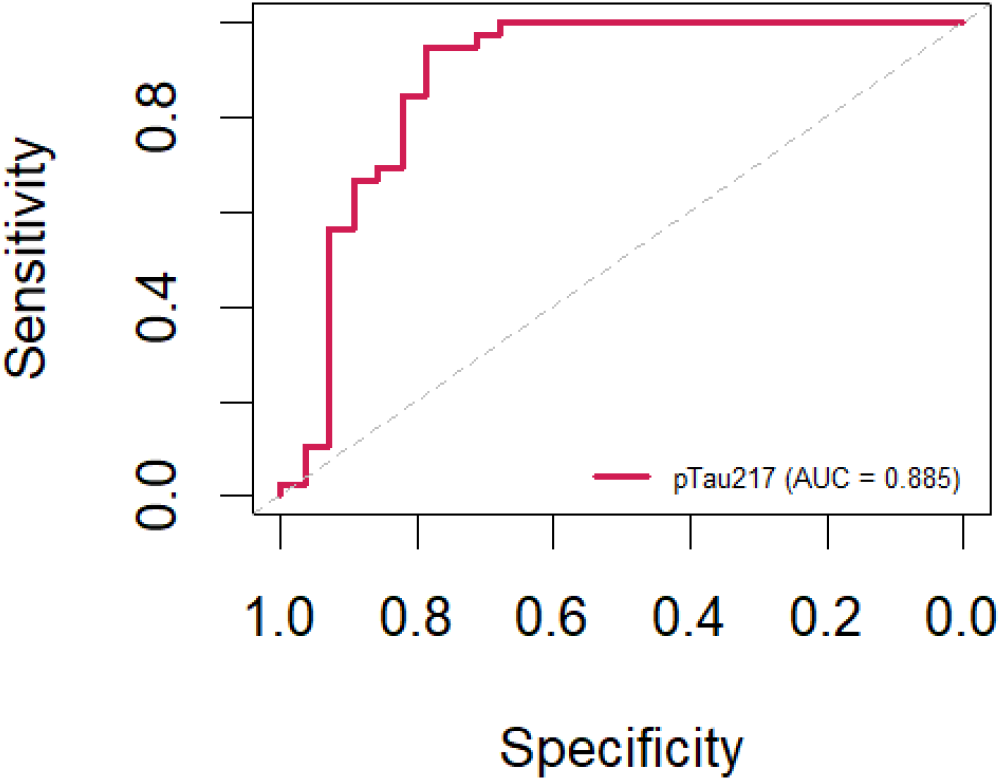
ROC analysis of p-tau217 biomarker levels.

## Discussion

This study presents a series of validation analyses of the PROTECT Cognitive Test System, which delivers eight computerised neuropsychological tests assessing domains of working memory, episodic memory, speed of attention, accuracy of attention and executive function. The outcomes show that the system can effectively discriminate between patients with dementia and healthy individuals and that it has excellent concurrent validity with the well validated plasma tau217 AD biomarker.

The comparison of performance between a dementia and healthy cohort with 9197 individuals shows that the PROTECT system identifies significant differences in performance in all tests across all cognitive domains of memory, attention and executive function. This is further strengthened by the correlation with the MoCA and the ROC analysis which shows excellent discriminative ability across all domains with highly significant AUC values of above 0.9 and both sensitivity and specificity of 90% for nearly all domains. This exceeds the generally accepted thresholds for clinical utility^27^.

Advances in biomarker technology have become an increasingly important and well validated part of the process for diagnosing AD and MCI due to AD, particularly for clinical trials and therapeutics focussing on amyloid or other key AD pathologies^28^. The correlation between a neuropsychological assessment and biomarker positivity is therefore an important potential metric of the value of a cognitive assessment battery. For this reason, the Pre-Clinical Alzheimer’s Cognitive Composite (PACC) was developed as a composite of cognitive measures that were strongly associated with amyloid positivity, and longitudinal outcome in amyloid-positive individuals^29^. However, the actual cross-sectional correlation between the PACC score and amyloid levels is much more modest (correlation coefficient R< 0.2)^30^. There are also data available for the correlations between scores on the widely used CogState computerized neuropsychology battery and AD biomarkers of tau and amyloid, with significant but modest correlations reported with correlation coefficients between 0.1 and 0.3^31^, but no evidence of significant correlations in a subsequent study examining neuroimaging correlates^32^. Within this study we examined the correlations of the PROTECT cognitive battery with plasma p-tau217, a biomarker which correlates highly with brain amyloid and tau pathology. Episodic memory, accuracy of attention and executive function as measured by the PROTECT cognitive system all had correlation coefficients >0.5 with plasma p-tau217 and a correlation of >0.4 was identified between working memory and the p-tau217 marker. This demonstrates a high level of correlation between the key PROTECT computerized neuropsychological tests and a well validated AD biomarker and suggests that the PROTECT cognitive test system has advantages over other current testing systems as a screening tool to identify participants who are biomarker positive for clinical trials and for treatment with disease-modifying anti-amyloid therapies.

This study provides robust data to support the use of the PROTECT cognitive test system for screening of individuals for early dementia risk in both healthcare and clinical trials, and highlights the potential for its use in combination with blood biomarkers. Its computerised, unsupervised delivery and track record for at-home delivery means it is scalable and accessible for the target patient group. Although the overall cohort size was substantial, further work is needed to validate the test system further in patients with dementia, further validate age specific thresholds and to confirm direct correlations with brain amyloid and tau pathology using PET neuroimaging.

## Acknowledgements

This paper represents independent research part-funded by the National Institute of Health Research (NIHR) Exeter Biomedical Research Centre and NIHR HealthTech Research Centre in Brain Health. The views expressed are those of the authors and not necessarily those of the NIHR or the Department of Health and Social Care. This paper was also supported by the NIHR Collaboration for Leadership in Applied Health Research and Care South-West Peninsula.

## Author Contributions

Anne Corbett: Conception and design, data analysis and interpretation, manuscript drafting and reviewing; Abbie Palmer: Data acquisition and analysis, manuscript reviewing; Millie Sander: Data acquisition and analysis, manuscript reviewing; Christine Davis: Design, data acquisition, manuscript reviewing; Kate Stych: Data acquisition, manuscript reviewing; Mary O’Leary: Data acquisition, manuscript reviewing; Jon Huntley: data interpretation, manuscript reviewing; Dag Aarsland: Conception and design, manuscript reviewing; Nicolas Castellano: Data acquisition, manuscript reviewing; Felipe Botero-Rodriguez: Data acquisition, manuscript reviewing; Gerard Griffieon: Data acquisition, manuscript reviewing; Mieke Nuytten: Data acquisition, manuscript reviewing; Nicholas Ashton: Data interpretation, manuscript reviewing; Hanna Huber: Data acquisition, manuscript reviewing; Adam Hampshire: Data acquisition, manuscript reviewing; Clive Ballard: Conception and design, data analysis and interpretation, manuscript drafting and reviewing.

## Data Availability

Data is available on request from the PROTECT study following approval. Requests can be directed to

